# Comparable 40 Hz Auditory Steady-State Responses in Children at Familial High Risk for Schizophrenia or Bipolar Disorder and Population-Based Controls

**DOI:** 10.1101/2025.04.11.25325628

**Authors:** Kit Melissa Larsen, Júlia Díaz-i-Calvete, Anna Hester Ver Loren van Themaat, Anne Amalie Elgaard Thorup, Kerstin Jessica Plessen, Merete Nordentoft, Hartwig Roman Siebner

## Abstract

**Background:** The 40-Hz auditory steady state response (ASSR) is reduced in patients with schizophrenia and bipolar disorder. Recent evidence suggests that ASSR may be altered in clinical high risk and familial high-risk populations. Here, we hypothesize that children aged 11-12 years with familial high risk for schizophrenia and bipolar disorder exhibit impairments in 40-Hz ASSR compared to population-based controls (PBC).

**Methods:** The cross-sectional study included 196 participants aged 11-12 years: 76 at familial high risk for schizophrenia (FHR-SZ), 50 at familial high risk for bipolar disorder (FHR-BP) and 70 PBC. ASSR power and inter-trial phase coherence (ITPC) were measured with 128-channel electroencephalography during regular and irregular 40-Hz auditory stimulation. Group differences in power and ITPC of the ASSR were assessed with independent linear mixed models for both the early and late ASSR component. Statistical analyses are reported with both frequentist p-values and Bayes factors.

**Results:** Both PBC and participants with FHR demonstrated stable ASSRs during regular 40 Hz click stimulation, but not during irregular. Bayes analyses showed moderate and anecdotal evidence against a difference in ASSR power or ITPC among the three groups in the early-latency and the late-latency component of the ASSR. Results remained unchanged after controlling for the presence of an Axis I disorder.

**Conclusions:** While reductions of 40-Hz ASSR are well-documented across psychiatric disorders and stages, our results show that these deficits are not evident in 11–12-year-old children at familial high risk for these disorders.

## 1 Introduction

Schizophrenia and bipolar disorder are severe mental disorders. A positive family history of either schizophrenia or bipolar disorder is the strongest risk factor for the development of these or other psychiatric disorders (1,2). Studying the offspring of affected parents can help identify abnormal developmental trajectories preceding clinical onset as well as factors that promote resilience despite a genetic risk. Both disorders involve widespread cortical dysfunctions and impaired integration and synchronization of neuronal activity across multiple brain regions (3–5). Studies into the mechanisms subserving synchronized brain function are critical to understand the neurodevelopment of schizophrenia and bipolar disorders.

Gamma-frequency rhythmic activity, including 40-Hz auditory steady-state responses (ASSR), is critical for coordinated brain function (6,7). ASSR can be non-invasively measured with electroencephalography (EEG), while participants listen to brief clicks or tones at a repetition rate of 40 Hz (8). It consists of two components: the early transient (0–100 ms) and the late-latency sustained “steady-state” component (200 ms – stimulus end) (9–11). The ability to generate 40-Hz ASSR relies on the balance between excitation and inhibition in neural networks (6), critically involving N-methyl-D-aspartate (NMDA) receptors and fast-spiking parvalbumin-positive GABAergic interneurons (10,12–14). A seminal study has shown that the ability to generate 40 Hz oscillations is increased in healthy participants when taken the GABA_A_ receptor– agonist lorazepam (15), emphasizing the role of GABA in the generation of 40-Hz ASSR.

Individuals with schizophrenia consistently show reduced 40-Hz ASSR (16), with good test-retest reliability in both healthy controls and patients (17). Similar reductions are observed in individuals with bipolar disorder, though with slightly smaller effect sizes (18). In schizophrenia, reductions are evident in early transient and late sustained ASSR components (19) which may vary depending on the illness stages (20). Despite the fact that data in bipolar disorder are more limited, similar patterns have been reported, particularly in relation to psychotic symptoms and in more severe psychopathology (21).

Impairments in the 40-Hz ASSR are not confined to chronic psychotic disorders, but are observed across various stages, including recent-onset of schizophrenia and first-episode psychosis (11,17), clinical high risk for psychosis (11,17,22,23) and unaffected first-degree relatives of schizophrenia patients (24). So far, it is unknown whether 40-Hz ASSR is affected in children with familial high risk of developing a psychiatric disorder. Following children with familial high risk of developing severe psychiatric disorders can help elucidating developmental aspects of the disorders. To fill this gap in the literature, our current study aims to provide an early cross-sectional evaluation of the 40-Hz ASSR in participants at age 11-12 years, born to parents with schizophrenia or bipolar disorder. We here provide results from the first wave while we are collecting multiple time points that will be published in separate longitudinal follow up studies. We hypothesized that children at familial high risk for severe psychiatric disorders would show a reduction in 40-Hz ASSR, particularly children of parents with schizophrenia. We further explored whether the presence of a lifetime Axis I diagnosis affects the ASSR.

## 2 Methods

### 2.1 Participants

We recruited 196 children from The Danish High Risk and Resilience Study (The VIA cohort) (25) that underwent a 40-Hz ASSR EEG assessment. The VIA study is a longitudinal register-based cohort study starting in 2012, when the participants were seven years of age (26). A total of 522 children were included. The EEG data presented here were collected in the first follow-up at age 11 years when neuroimaging modalities were included in the assessment (25). Of the 196 children participating in this sub study, 76 had at least one parent with a diagnosis of schizophrenia (FHR-SZ), 50 had at least one parent diagnosed with bipolar disorder (FHR-BP) and finally 70 had parents without these disorders, referred to as population-based controls (PBC). The EEG data were collected at the Danish Research Centre for Magnetic Resonance (DRCMR), Copenhagen University Hospital Hvidovre. The study was approved by the National Committee on Health Research Ethics (Protocol number: H 16043682) and the Danish Data Protection Agency (ID number RHP-2017-003, I-suite no. 05333) and conducted in accordance with the Declaration of Helsinki.

### 2.2 Clinical variables

We used the Child Behavior Checklist (CBCL) school-age version to assess behavioral problems (27) and the Children’s Global Assessment Scale (CGAS) to assess the level of general functioning throughout the previous month (28). Current or past presence of any Axis I disorder were identified through the semi-structured interview for Affective Disorders and Schizophrenia for School-Age Children-Present and Lifetime Version (K-SADS-PL) (29). According to the procedure (30), none of the participants in the present study met criteria for Axis I psychotic disorder (DSM IV 298.9/298.8/297.1/292.30/295.90) at inclusion in this sub study of VIA. Group differences in age, CBCL and CGAS were assessed using one-way analysis of variance. Group differences in sex and presence of Axis I Disorder were assessed using a Pearson’s Chi-squared test. Drop-out analyses were performed using paired t-tests between the groups on CBCL scores.

### 2.3 ASSR paradigm

To evoke steady-state gamma activity, participants were presented with a train of short clicks delivered regularly at a mean click-repetition frequency of 40 Hz, as in Larsen et. al (2018) (23). Each click lasted 1 ms followed by a constant inter-click interval of 25 ms (i.e., regular 40 Hz train) or randomly jittered between 11 and 37 ms while maintaining a mean click frequency of 40 Hz (i.e., irregular 40 Hz train). Each click train was 1 s, followed by a pause of 2 s, resulting in a stimulus onset asynchrony of 3 s. The temporal structure of the irregular click train condition was kept constant within subjects but changed randomly across subjects. With this irregular condition that is matched to the regular condition on acoustic features, we could control whether the temporal regularity of the 40 Hz click train was critical to evoke abnormal auditory cortical processing restricted to ASSR. The stimuli were delivered binaurally via insert-earphones at a sound pressure level of 85 dB (E-ARTONE 3A), with the MatLab-based PsychToolbox as presentation software using an external soundcard (RME Babyface 22-Channel, 192 kHz Bus-powered, Haimhausen, Germany). Participants were seated in a comfortable chair, were instructed to relax and to constantly look at a fixation cross on the screen in front of them without paying particular attention to the sounds. We applied the regular and irregular 40 Hz trains in two separate runs consisting of 120 trials of each condition. A single run lasted 6 min.

### 2.4 EEG Recordings and Preprocessing

EEG data were recorded using a 128 channel BioSemi ActiveTwo System (BioSemi, Amsterdam, Netherlands) at 4096 Hz of sampling frequency. All offline preprocessing was performed using SPM12 (http://www.fil.ion.ucl.ac.uk/spm/) running in MatLab. Preprocessing steps included high- and low-pass filtering with a 5th order Butterworth filter with a cut-off of 0.1 Hz and 80 Hz respectively. Additionally, a notch filter was applied [48 52] to attenuate 50 Hz power line noise and data were downsampled to 512 Hz. The signals were referenced to the average of the signal from the two mastoid channels. Data were epoched with a peristimulus interval of−1000 ms to 2000 ms. Finally, artefact rejection was performed using a simple threshold technique rejecting trials if amplitudes exceeded ±130 μV, with channels marked as bad if more than 70% trials were rejected. On average the rejected number of trials did not differ between groups (p = 0.879).

Of the included 196 participants 11 participants (three FHR-SZ, one FHR-BP, seven PBC) had to be excluded due to too noisy data or a problem with the trigger registration during data collection. Hence, a total of 185 participants were included in the final analyses. Of these, nine individuals were included in the analysis but did not have both conditions available: five were missing the regular condition and four the irregular condition. The demographics of the sample were assessed on these 185 participants (see Table 1).

**Table 1:** Demographic data for the cohort. *CBCL: child behavior check list, CGAS: Children’s Global Assessment Scale*. Group differences in sex and presence of Axis I Disorder were assessed using a Pearson’s Chi-squared test. Group differences in age, CBCL and CGAS were assessed using a one-way analysis of variance.

|  | Total | FHR-SZ | FHR-BP | PBC | p-value |
| --- | --- | --- | --- | --- | --- |
| Children, N | 185 | 73 | 49 | 63 | - |
| Females, N (%) | 91 (49) | 35 (48) | 24 (49) | 32 (51) | 0.946 |
| Age, mean (SD) | 12.13 (0.29) | 12.13 (0.29) | 12.14 (0.27) | 12.13 (0.31) | 0.897 |
| **CBCL, mean (SD)** |  |  |  |  |  |
| CBCL, Total | 17.50 (17.44) | 21.30 (20.41) | 16.76 (15.42) | 13.67 (14.27) | 0.040 |
| CBCL, internalizing | 5.79 (5.97) | 6.39 (6.42) | 5.65 (4.98) | 5.19 (6.17) | 0.510 |
| CBCL, externalizing | 3.98 (5.44) | 5.25 (6.94) | 3.86 (4.59) | 2.59 (3.46) | 0.019 |
| CGAS | 70.54 (15.31) | 66.23 (15.19) | 70.51 (15.24) | 75.65 (14.11) | 0.002 |
| Lifetime Axis I disorder, excl. elimination disorders, N (%) | 78 (42) | 37 (51) | 22 (45) | 19 (30) | 0.050 |

### 2.5 Time-Frequency Analysis

The epoched data were wavelet-transformed using a Complex Morlet wavelet with 12 cycles and frequency band of interest covering frequencies from 2 to 60 Hz at 1 Hz resolution. Power and inter-trial phase coherence (ITPC) were extracted from the wavelet coefficients. The power was rescaled relative to the baseline using the time window –800 ms to –10 ms and presented in dB. The amplitude of the ITPC reflects the phase consistency across trials for a given channel, time, and frequency point, bounded between 0 to 1. A value of 1 reflects perfect phase synchrony across trials. To compare power and ITPC values for the different frequency bands, values were extracted during the time of stimulation from 0-100 ms for the early latency and 300 – 1000 ms for the late latency in the frequency range [38 42]. This extraction of values was performed for electrode Cz and the 5 electrodes surrounding Cz and a mean value of these 6 channels are presented. The Cz and surrounding electrodes show the highest topographical signal for 40-Hz ASSR (31).

### 2.6 Assessing Group Differences in ASSR responses

Group differences in power and ITPC of the ASSR were assessed with independent linear mixed models for both the early latency ASSR and the late latency ASSR, resulting in four models. Age was included as a covariate in all four models, given the sensitive period regarding brain development (32) with a potential influence in the maturation of auditory responses (33). Sex was further added as a covariate since differences are frequently reported during brain development (34). Thus, the models accounted for independent variables of group (factor of 3 levels: FHR-SZ, FHR-BP, PBC), age, sex and within-subject ASSR stimuli condition (factor of 2 levels: regular, irregular). The participants’ ID was used as a clustering variable to control for repeated measures within-subject and between-subject variability (model with random intercept). We were interested in investigating if group effects differed by condition. We therefore included the interaction term group-by-condition. If no significant interaction effect was detected, the interaction term was removed to assess the effect of group independently. We report both frequentist p-values as well as Bayes factors (BF_10_). All statistical analyses were performed in R (version 4.4.0, 2024-04-24) using the packages BayesFactor, lme4 and lmerTest. When needed, Bonferroni was used to correct for multiple comparison.

## 3 Results

At the time of examination, the children had a mean age of 12.13 years, and the three groups were comparable in both age and sex (see Table 1). Children at FHR showed higher CBCL total score and externalizing scores compared to PBC, indicating higher ratings of behavioral problems in the FHR group. CBCL internalizing score did not differ between groups. The level of global functioning as measured with CGAS was higher in PBC compared to the two FHR groups (p = 0.002). The presence of any lifetime Axis I diagnosis did not significantly differ between groups. Table 1 lists the summary statistics for each group.

Drop-out analyses revealed no difference in CBCL scores in participants that took part of this EEG sub study compared to those that did not, see supplementary material for details.

### 3.1 Group differences in the ASSR

PBC showed a clearly discernible ASSR around 40 Hz which was temporally confined to the time of regular click stimulation. As expected, jittering the time distribution of the clicks along the second of 40 Hz stimulation, as evident in the irregular condition plots, clearly diminished the response (Figures 1 and 2). Visual inspection of the time-frequency plots indicated that the FHR-SZ group has a more pronounced response around 40 Hz when compared to PBC, whereas FHR-BP had a weakened response at the group level. The response to the irregular condition were comparable between groups (Figure 2).

**Figure 1.**
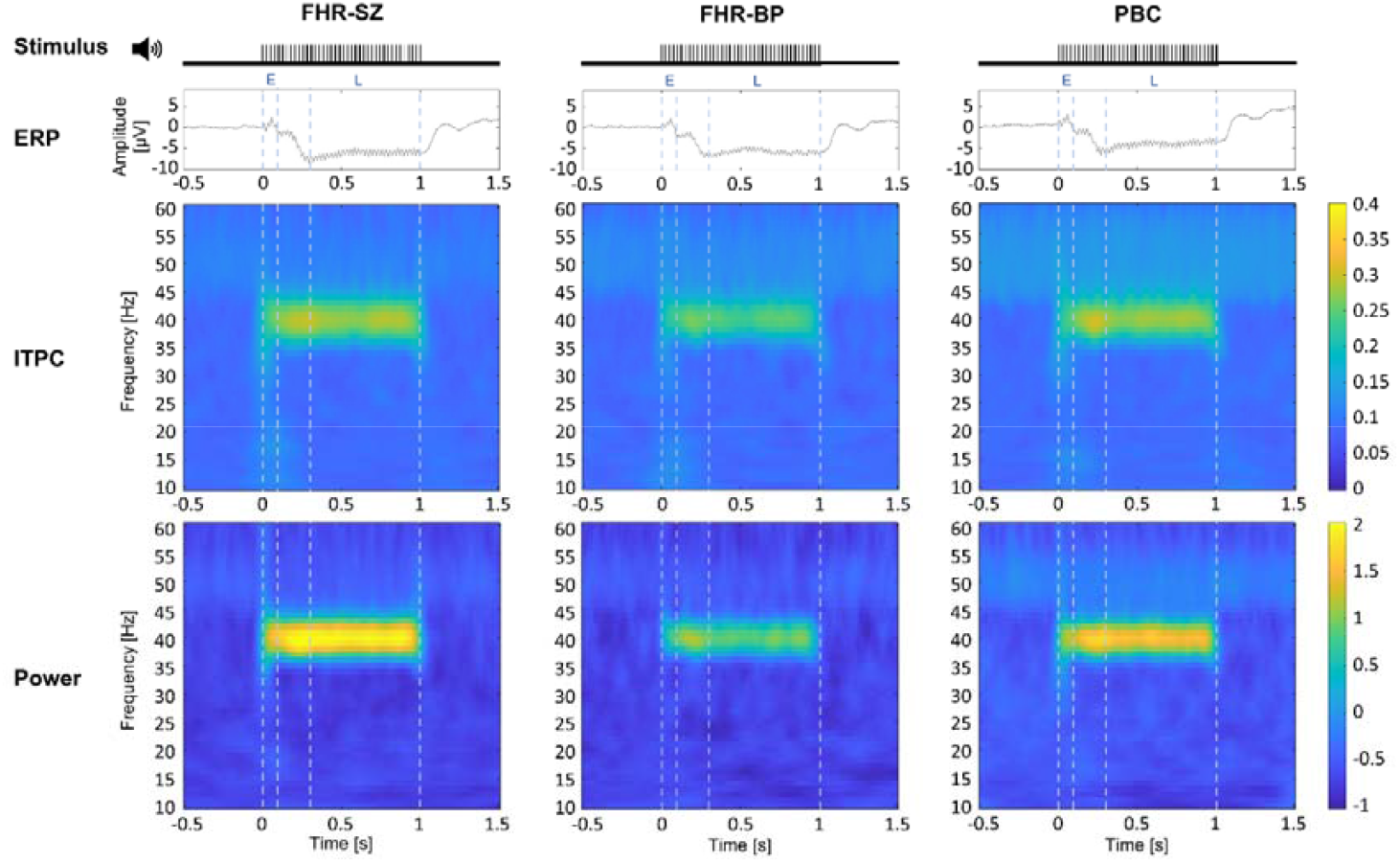
Time-frequency responses for the regular condition shown for all three groups as grand mean across participants; FHR-SZ left, FHR-BP middle and PBC right. Time frequency responses are shown as a mean of the responses from electrode Cz and the 5 electrodes surrounding Cz. At the top, the regular stimulation is visualized followed by the ERP responses, ITPC and power respectively. The dotted vertical lines indicate the time-bins used for the analyses of the early and late ASSR respectively.

**Figure 2.**
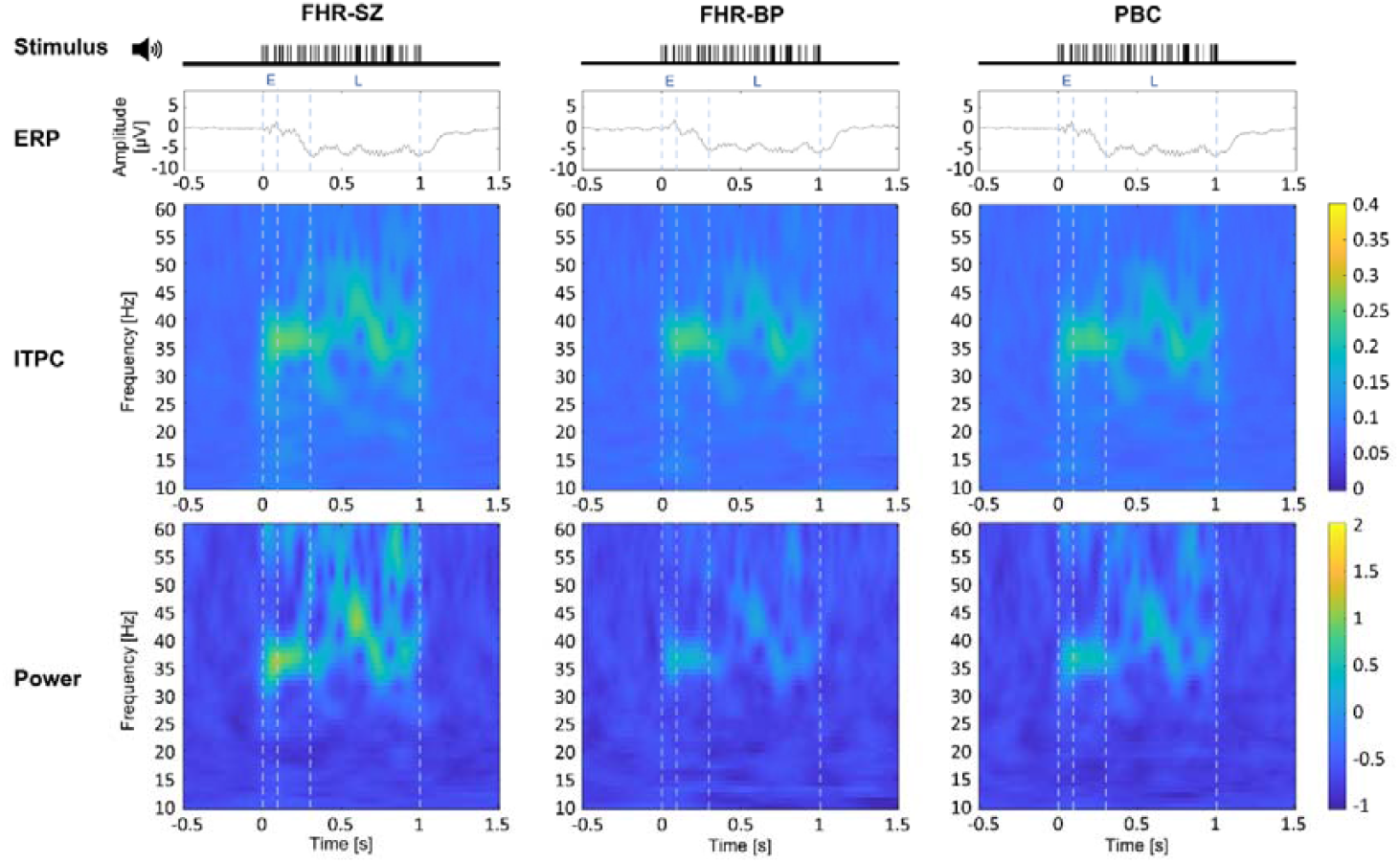
Time-frequency responses for the irregular condition shown for all three groups as grand mean across participants; FHR-SZ left, FHR-BP middle and PBC right. Time frequency responses are shown as a mean of the responses from electrode Cz and the 5 electrodes surrounding Cz. At the top, the regular stimulation is visualized followed by the ERP responses, ITPC and power respectively. The dotted vertical lines indicate the time-bins used for the analyses of the early and late ASSR respectively.

#### 3.1.1. Early-latency transient response

Results from the linear mixed models for the transient response showed that the regular condition was overall more efficient in evoking the 40-Hz ASSR than the irregular condition across groups. This was the case for both ITPC and power (ITPC: *F*(1,178.13) = 62.953, *p* < 0.001, BF_10_ > 1000, power: *F*(1,181.17) = 34.988, *p* < 0.001, BF_10_ = 612.959) (see Figure 3). Both power and ITPC were similar between groups (ITPC: *F*(2,181.95) = 0.872, *p* = 0.420, BF_10_ = 0.102, power: *F*(2,182.94) = 1.302, *p* = 0.275, BF_10_ = 0.173), with the Bayes factor showing anecdotal evidence of no effect. No effect of age or sex was observed.

**Figure 3.**
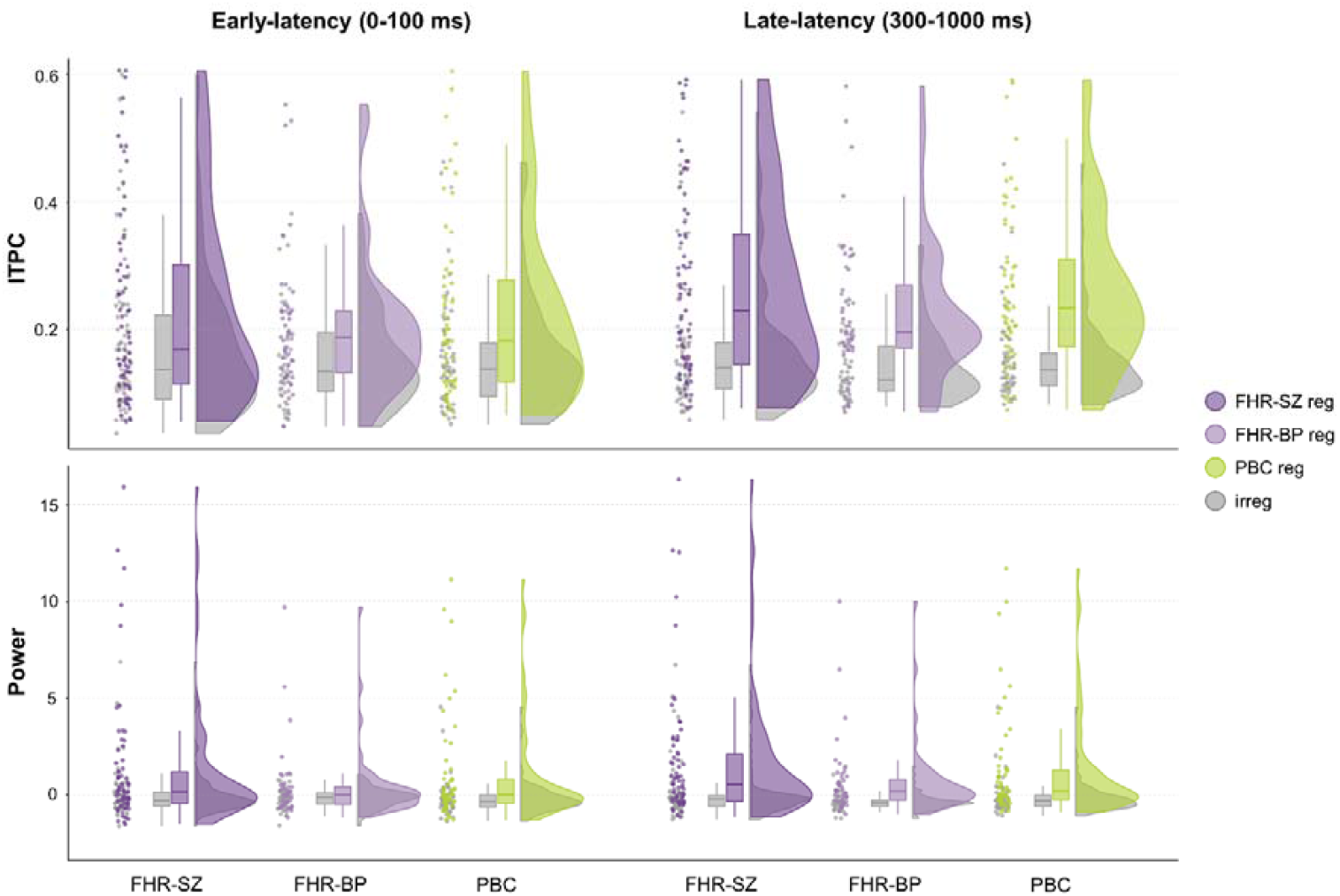
Distribution of extracted ITPC and power values (regular condition) for FHR-SZ (dark purple), FHR-BP (light purple) and PBC (green). Each data point represents a participant. Values for the irregular condition are shown in grey next to the corresponding regular condition. Values for the early-latency (0-100 ms) are shown to the left and values for the late-latency (300-1000 ms) are shown to the right.

#### 3.1.2 Late-latency steady-state response

For the steady-state response, the regular condition as expected was more efficient to evoke the 40 Hz gamma entrainment compared to the irregular condition for both power and ITPC (ITPC: F(1,177.28) = 212.896, p < 0.001, BF_10_ > 1000, power: *F*(1,180.00) = 80.288, *p* < 0.001, BF_10_ > 1000), see Figure 3. Like the early transient response, the late steady-state responses were comparable between groups (ITPC: *F*(2,181.96) = 1.873, *p* = 0.157, BF_10_ = 0.472, power: *F*(2,183.03) = 2.204, *p* = 0.113, BF_10_ = 0.621), again with the Bayes factor showing anecdotal evidence of no effect. Likewise, age and sex showed no effect on the steady state responses.

### 3.2 Effect of lifetime Axis I diagnosis

We wished to examine whether the presence of any lifetime Axis I diagnosis would influence the ASSR. We therefore performed a post-hoc analysis, adding the presence of an Axis I diagnosis (as binary variable) as a factor to the models. In neither of the models, the presence of a lifetime Axis I diagnosis had an effect or changed the observed results.

## 4 Discussion

This study provides novel evidence that the 40-Hz ASSR in 11–12-year-old children at familial high risk for schizophrenia and bipolar disorder are comparable to population-based controls of the same age. Importantly, controlling for the presence of any past or present Axis I diagnosis revealed no significant effect on 40-Hz gamma-band ASSR in this age range. While reductions of 40-Hz ASSR are well-documented across psychiatric disorders and stages (21,22,34,35), our negative results show that these deficits are not evident in children at familial high risk for these disorders at stages around the onset of puberty. Thus, these results contribute to the neurodevelopmental understanding of these conditions and may help identify the critical time window for investigating the underlying maturation processes potentially impaired.

Previous studies report small, often non-significant, reductions in 40-Hz ASSR power in individuals at risk for psychosis (35), contrasting with moderate-to-large effects in both power and phase in first-episode psychosis patients (35). Consistent with this, the absence of psychotic disorders in our current study at age 11, suggests conserved auditory processing in FHR-SZ and FHR-BP children prior to the onset of schizophrenia or bipolar spectrum disorders. In fact, our results gave anecdotal to moderate evidence against a significant difference. Here, we further showed that the presence of an Axis I diagnoses did not have effect on the ASSR in the current study, which is known to be elevated in high risk groups (30,36). However, this finding does not exclude the general influence of comorbidities on the ASSR.

Gamma oscillations follow a developmental trajectory that peaks in mid-adolescence, increasing steadily from ages 8–10 through 14–16 (37). This maturational trajectory is thought to involve the development of parvalbumin positive fast-spiking basket cells and GABAergic inhibition within interneuron circuits (37). Reduced NMDA-mediated excitation of fast-spiking interneurons may also enhance gamma activity and ASSR by disinhibiting pyramidal cells (37). In individuals predisposed to psychosis-spectrum disorders, disruptions in these maturational processes could cause deviations in gamma activity trajectories, which may emerge later in adolescence. Given that the conversion rate to schizophrenia or bipolar disorder in familial high-risk (FHR) cohorts is estimated at 5–8% (2), most included children are unlikely to develop these disorders. As a result, large effect sizes are needed to detect significant group differences. Future longitudinal assessments in this cohort are essential to track developmental changes in gamma synchrony and identify potential divergences related to risk.

To our knowledge, this is the first study on 40-Hz ASSR including both schizophrenia and bipolar high-risk groups at the same age in early adolescence. Replications are warranted to confirm the absence of significant group differences in evoked gamma entrainment at this age. A key aspect of the present study is that children were categorized based on their parents’ psychiatric history. However, children of parents with severe mental disorders face an elevated risk not only of developing the specific disorder as their parent has, but also a broader spectrum of psychiatric conditions (2,38). Since this cohort is being followed into adulthood, we will be able to use this information later in the follow up studies. Having the future outcomes and potential diagnoses will allow to trace back on these ASSR data and correlate the measurements with both neuroimaging and phenotyping data about the participants. This could contribute to personalized approaches for risk assessments and biometric identification.

While our study has several strengths, it also has limitations. We have provided a cross-sectional snapshot from the early phase, prior to the clinical onset of severe mental disorders. To draw definitive conclusions, longitudinal data are needed to assess individual trajectories. Given that the overall VIA study employs a longitudinal design, we will have future data to track the development of the 40-Hz ASSR in these children. Although this represents a limitation in the current study, the findings at ages 11-12 years still offer a valuable perspective on the early stages before clinical manifestation.

In conclusion, our findings indicate comparable responses of the 40-Hz ASSR in children at FHR for schizophrenia and bipolar disorder to population-based controls. Given that reductions in 40-Hz ASSR may not be evident in at-risk children prior to onset, this could prove valuable in clarifying the neural mechanisms underlying the onset and progression of psychotic episodes, as well as the developmental timing of gamma disturbances in affected individuals. Our negative finding highlights the critical importance of longitudinal studies on children at high risk for psychiatric disorders. While the study is cross-sectional, longitudinal data are currently collected and will provide opportunities of correlating different functional responses and structural brain characteristics longitudinally, covering the vulnerable period of adolescence and beyond in the absent of psychotic medications.

## Supporting information

Drop-out analyses

## Data Availability

Data is not available due to Danish law

## Acknowledgements

The current study received funding from the Research Fund in the Capital Region of Denmark, The Lundbeck Foundation Initiative for Integrative Psychiatric Research (iPSYCH R102-A9118), the Mental Health Services of the Capital Region of Denmark and Innovation Fund Denmark (6152-00002B). KML received funding from The Lundbeck Foundation (R322-2019-2311). Hartwig Roman Siebner received funding from Innovation Fund Denmark (9068-00025B) and from The Lundbeck Foundation (R336-2020-1035).

Thanks to all families and children for their time invested in the Danish High Risk and resilience Study. For assistance in data collection, we would like to thank Benthe Emke Vink, Daban Khalid Ameen Sulaiman, Line Korsgaard Johnsen, and Jonas Schultz Ingerslev and Jessica Ohland for data management. Further, we would like to thank Nanna Ørslev Weye for help with the drop-out analyses. Finally, we would like to thank all staff involved in the Danish High Risk and Resilience study.

## Author contribution

KML: data curation, formal analyses, visualization and writing and editing of the paper. JDC: formal analyses, data curation, visualization, and writing. HVLT: project administration, investigation and editing of the paper. AAET, KJP and MN: conceptualization, funding acquisition, supervision as well as editing the manuscript. HRS: conceptualization, funding acquisition, supervision as well as writing and editing the manuscript.

## References

1. Hallmayer J (2000): The epidemiology of the genetic liability for schizophrenia. Aust N Z J Psychiatry 34 Suppl: S47–S55.

2. Uher R, Pavlova B, Radua J, Provenzani U, Najafi S, Fortea L, et al. (2023): Transdiagnostic risk of mental disorders in offspring of affected parents: a meta-analysis of family high-risk and registry studies. World Psychiatry 22: 433–448.

3. Uhlhaas PJ, Singer W (2010): Abnormal neural oscillations and synchrony in schizophrenia. Nat Rev Neurosci 11: 100–113.

4. Friston K, Brown HR, Siemerkus J, Stephan KE (2016): The dysconnection hypothesis (2016). Schizophr Res 176: 83–94.

5. Stephan KE, Baldeweg T, Friston KJ (2006): Synaptic plasticity and dysconnection in schizophrenia. Biol Psychiatry 59: 929–39.

6. Buzsáki G, Wang X-J (2012): Mechanisms of gamma oscillations. Annu Rev Neurosci 35: 203–25.

7. Singer W (2018): Neuronal oscillations: unavoidable and useful? Eur J Neurosci 48: 2389–2398.

8. Plourde G, Stapells DR, Picton TW (1991): The Human Auditory Steady-State Evoked Potentials. Acta Otolaryngol 111: 153–160.

9. Roß B, Picton TW, Pantev C (2002): Temporal integration in the human auditory cortex as represented by the development of the steady-state magnetic field. Hear Res 165: 68–84.

10. Grent-’t-Jong T, Brickwedde M, Metzner C, Uhlhaas PJ (2023): 40-Hz Auditory Steady-State Responses in Schizophrenia: Toward a Mechanistic Biomarker for Circuit Dysfunctions and Early Detection and Diagnosis. Biol Psychiatry 94: 550–560.

11. Tada M, Kirihara K, Koshiyama D, Nagai T, Fujiouka M, Usui K, et al. (2023): Alterations of auditory-evoked gamma oscillations are more pronounced than alterations of spontaneous power of gamma oscillation in early stages of schizophrenia. Transl Psychiatry 13. 10.1038/S41398-023-02511-5

12. Bartos M, Vida I, Jonas P (2007): Synaptic mechanisms of synchronized gamma oscillations in inhibitory interneuron networks. Nat Rev Neurosci 8: 45–56.

13. Sohal VS, Zhang F, Yizhar O, Deisseroth K (2009): Parvalbumin neurons and gamma rhythms enhance cortical circuit performance. Nature 459: 698–702.

14. Carlén M, Meletis K, Siegle JH, Cardin JA, Futai K, Vierling-Claassen D, et al. (2012): A critical role for NMDA receptors in parvalbumin interneurons for gamma rhythm induction and behavior. Mol Psychiatry 17: 537–548.

15. Toso A, Wermuth AP, Arazi A, Braun A, Jong TG-’t, Uhlhaas PJ, Donner TH (2024): 40 Hz Steady-State Response in Human Auditory Cortex Is Shaped by Gabaergic Neuronal Inhibition. J Neurosci 44: e2029232024.

16. Thuné H, Recasens M, Uhlhaas PJ (2016): The 40-Hz Auditory Steady-State Response in Patients With Schizophrenia: A Meta-analysis. JAMA psychiatry. 10.1001/jamapsychiatry.2016.2619

17. Roach BJ, D’Souza DC, Ford JM, Mathalon DH (2019): Test-retest reliability of time-frequency measures of auditory steady-state responses in patients with schizophrenia and healthy controls. NeuroImage Clin 23. 10.1016/J.NICL.2019.101878

18. Jefsen OH, Shtyrov Y, Larsen KM, Dietz MJ (2022): The 40-Hz auditory steady-state response in bipolar disorder: A meta-analysis. Clin Neurophysiol 141: 53–61.

19. Griskova-Bulanova I, Hubl D, van Swam C, Dierks T, Koenig T (2016): Early- and late-latency gamma auditory steady-state response in schizophrenia during closed eyes: Does hallucination status matter? Clin Neurophysiol 127: 2214–2221.

20. Tada M, Nagai T, Kirihara K, Koike S, Suga M, Araki T, et al. (2014): Differential Alterations of Auditory Gamma Oscillatory Responses Between Pre-onset High-risk Individuals and First-episode Schizophrenia. Cereb Cortex 26: 1027–35.

21. Parker DA, Hamm JP, McDowell JE, Keedy SK, Gershon ES, Ivleva EI, et al. (2019): Auditory steady-state EEG response across the schizo-bipolar spectrum. Schizophr Res 209: 218–226.

22. Wang J, Li J, Tang Y, Liu X, Qian Z, Zhang T, et al. (2024): Impaired 40-Hz and intact hierarchical organization mode of auditory steady-state responses among individuals with clinical high-risk for psychosis. Prog Neuropsychopharmacol Biol Psychiatry 135. 10.1016/J.PNPBP.2024.111123

23. Larsen KM, Pellegrino G, Birknow MR, Kjær TN, Baaré WFC, Didriksen M, et al. (2018): 22q11.2 Deletion Syndrome Is Associated With Impaired Auditory Steady-State Gamma Response. Schizophr Bull. 10.1093/schbul/sbx058

24. Rass O, Forsyth JK, Krishnan GP, Hetrick WP, Klaunig MJ, Breier A, et al. (2012): Auditory steady state response in the schizophrenia, first-degree relatives, and schizotypal personality disorder. Schizophr Res 136: 143–149.

25. Thorup AAE, Hemager N, Søndergaard A, Gregersen M, Prøsch ÅK, Krantz MF, et al. (2018): The Danish High Risk and Resilience Study—VIA 11: Study Protocol for the First Follow-Up of the VIA 7 Cohort −522 Children Born to Parents With Schizophrenia Spectrum Disorders or Bipolar Disorder and Controls Being Re-examined for the First Time at Age 1. Front Psychiatry 9. 10.3389/fpsyt.2018.00661

26. Thorup AAE, Jepsen JR, Ellersgaard DV, Burton BK, Christiani CJ, Hemager N, et al. (2015): The Danish High Risk and Resilience Study – VIA 7 - a cohort study of 520 7-year-old children born of parents diagnosed with either schizophrenia, bipolar disorder or neither of these two mental disorders. BMC Psychiatry 15: 233.

27. Achenbach, T. M., & Edelbrock C (1983): Manual for the Child Behavior Checklist and Revised Child Behavior Profile.

28. Shaffer D, Gould MS, Brasic J, Fisher P, Aluwahlia S, Bird H (1983): A children’s global assessment scale (CGAS). Arch Gen Psychiatry 40: 1228–1231.

29. Kaufman J, Birmaher B, Brent D, Rao U, Flynn C, Moreci P, et al. (1997): Schedule for Affective Disorders and Schizophrenia for School-Age Children-Present and Lifetime Version (K-SADS-PL): initial reliability and validity data. J Am Acad Child Adolesc Psychiatry 36: 980–988.

30. Gregersen M, Søndergaard A, Brandt JM, Ellersgaard D, Rohd SB, Hjorthøj C, et al. (2021): Mental disorders in preadolescent children at familial high-risk of schizophrenia or bipolar disorder - a four-year follow-up study: The Danish High Risk and Resilience Study, VIA 11. J Child Psychol Psychiatry. 10.1111/JCPP.13548

31. Başar E (2013): A review of gamma oscillations in healthy subjects and in cognitive impairment. Int J Psychophysiol 90: 99–117.

32. Gogtay N, Giedd JN, Lusk L, Hayashi KM, Greenstein D, Vaituzis AC, et al. (2004): Dynamic mapping of human cortical development during childhood through early adulthood. Proc Natl Acad Sci U S A 101: 8174–8179.

33. Corcoran CM, Stoops A, Lee M, Martinez A, Sehatpour P, Dias EC, Javitt DC (2018): Developmental trajectory of mismatch negativity and visual event-related potentials in healthy controls: implications for neurodevelopmental vs. neurodegenerative models of schizophrenia. Schizophr Res 191: 101.

34. Kaczkurkin AN, Raznahan A, Satterthwaite TD (2018): Sex differences in the developing brain: insights from multimodal neuroimaging. Neuropsychopharmacol 2018 441 44: 71–85.

35. Zouaoui I, Dumais A, Lavoie ME, Potvin S (2023): Auditory Steady-State Responses in Schizophrenia: An Updated Meta-Analysis. Brain Sci 13: 1722.

36. Keshavan M, Montrose DM, Rajarethinam R, Diwadkar V, Prasad K, Sweeney JA (2008): Psychopathology among offspring of parents with schizophrenia: relationship to premorbid impairments. Schizophr Res 103: 114–120.

37. Cho RY, Walker CP, Polizzotto NR, Wozny TA, Fissell C, Chen CMA, Lewis DA (2015): Development of sensory gamma oscillations and cross-frequency coupling from childhood to early adulthood. Cereb Cortex 25: 1509–1518.

38. Rasic D, Hajek T, Alda M, Uher R (2014): Risk of Mental Illness in Offspring of Parents With Schizophrenia, Bipolar Disorder, and Major Depressive Disorder: A Meta-Analysis of Family High-Risk Studies. Schizophr Bull 40: 28–38.

