## Supplementary material for "Comparable 40 Hz Auditory Steady-State Responses in Children at Familial High Risk for Schizophrenia or Bipolar Disorder and Population-Based Controls": Drop-out analyses

Drop-out analyses comparing CBCL values for the participants that took part of the EEG study versus those that did not take part of the EEG sub-study. All values here, are from the VIA7 cohort, to show representativeness from the original cohort.

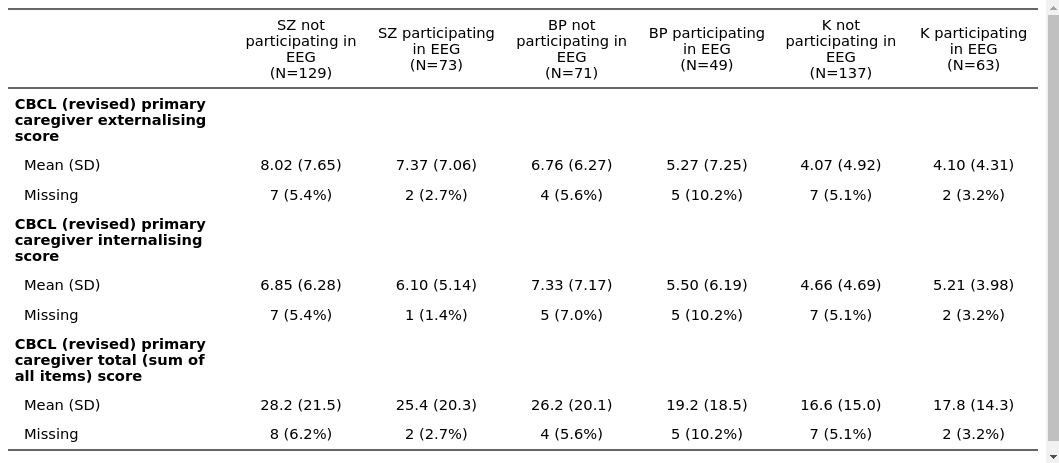

P-values from pair-wise comparisons of CBCL scores of participants participating in EEG vs those in VIA that did not take part in the EEG study.

|  | SZ in EEG vs SZ not in EEG | BP in EEG vs BP not in EEG | PBC in EEG vs PBC not in EEG |
| --- | --- | --- | --- |
| CBCL total | p = 0.311 | p = 0.053 | p = 0.679 |
| CBCL internalizing | p = 0.366 | p = 0.094 | p = 0.527 |
| CBCL externalizing | p = 0.487 | p = 0.227 | p = 0.976 |
